# ACUTE KIDNEY INJURY AS A PREDICTOR OF IN-HOSPITAL MORTALITY IN PATIENTS WITH A DIAGNOSIS OF SEVERE COVID-19

**DOI:** 10.1101/2024.05.31.24308297

**Authors:** Anthonela Cruz Ordinola, Carlos Fajardo Arriola, Raúl Sandoval Ato

## Abstract

**OBJECTIVE:** Determine if Acute Kidney Injury (AKI) is a predictor of In-hospital Mortality in severe Covid-19 patients.

**METHODS:** A retrospective cohort study was carried out with severe Covid-19 patients from the Hospital de la Amistad Perú - Corea Santa Rosa II - Piura, using a multivariate analysis with the bivariate logistic regression technique and the method = Advance by steps (ratio of likelihood), to calculate a predictive model for Mortality in Covid-19 patients exposed to ARI.

**RESULTS:** Mortality was higher in the cohort exposed to ARI (95% vs 71.9%) and a 5-fold RR of death in the patient exposed to ARI compared to the Covid-19 patient not exposed to ARI.

**CONCLUSION:** Exposure to ARI increases the risk of mortality by 5 times, increasing the risk of mortality based on the increase in the SOFA score upon admission to the ICU and increase in mg/dl of C-Reactive Protein.

## INTRODUCTION

Covid-19 is a pathology characterized by being very contagious, infectious and mainly pneumonic. The WHO considered this disease a pandemic that has great implications for public health throughout the planet.^1-2^ Among the main clinical manifestations are respiratory symptoms such as cough, fever and fatigue, which triggers, in its most severe forms, a respiratory distress syndrome.^3-4^ Furthermore, it has been proven that this type of serious viral infections cause multiple organ dysfunction, and can affect not only the lungs but also cause kidney damage.^5-6^

Patients with Covid-19 present clinical deterioration that develops rapidly as the main complications, and it was observed that a group of patients developed acute renal dysfunction.^7-8^ Currently, the cause of acute kidney injury in Covid-19 is the subject of study, for which various mechanisms have been described that explain kidney involvement. One of them explains that the virus has a high affinity for ACE2 (angiotensin-converting enzyme 2 receptor), which is expressed both in lung tissue and in kidney cells, being expressed 100 times higher in kidneys than in the lung.^9-10^

ARF has a great impact on the course of various diseases, being accompanied by high morbidity, being considered a very important issue, requiring specialized interventions for this reason. However, ARF during COVID-19 disease can present in a different way, where patients with mild kidney injury may go unnoticed, which is why ARF should be considered primarily as a risk factor.^11-12^

It has been described in various studies that among patients diagnosed with Covid-19 there is an association between sepsis and death from Covid-19, this mainly as a result of direct infection by Sars-Cov-2 causing damage to multiple organs including the kidney, potentially trigger ARF and be an indicator of multiorgan failure, defined by the SOFA score, this being an excellent diagnostic marker. As has been well demonstrated, sepsis is generally a complication of bacterial infections, however, viral infections could also trigger sepsis, as evidenced in Covid-19 disease.^13^

To date, numerous cases of Covid-19 (>4 million) have been registered in Perú with a mortality rate of 4.87% at the national level, and with a fatality percentage of 7.29% in our region of Piura (more than 13 thousand deaths)^14^, being the cause of more hospitalizations in the ICU. Epidemiological data show us that the highest mortality rate was recorded between the years 2020 and 2021^14^ where the Covid-19 pandemic was developing for the first time in Perú and therefore the population was not yet immunized, being more vulnerable. to trigger severe forms of the disease. Furthermore, various studies carried out in Perú have revealed that ARI related to Covid-19 disease is synonymous with severity and can trigger death in these patients.^15^

This study aims to identify factors that predict a negative result taking into account that currently there is no longer a high incidence of severe cases of Covid-19, due to the various vaccination schedules that were established, however, there are still patients who could not adequately comply with the vaccination against Covid-19 and could develop severe forms of the disease and subsequently trigger ARI, thus increasing the risk of mortality.

Therefore, this research seeks to determine if ARF is a predictor of In-hospital Mortality in severe Covid-19 patients.

## METHODS

### DESIGN AND SAMPLE

This study has a Retrospective Cohort design. We included 200 patients >18 years old diagnosed with severe Covid-19 from Hospital de la Amistad Perú - Corea Santa Rosa II - Piura (HAPCII-2) from 04/01/2020 to 03/30/2021. Patients were included with creatinine tests, evaluating Glomerular Filtration values to determine if they presented ARF and their corresponding stage, also including patients with comorbidities such as Type 2 DM, HTN, Obesity, Sepsis and Septic Shock recorded in the medical records. Patients with CKD stage 4-5, on hemodialysis and without creatinine tests on admission were excluded.

### DATA COLLECTION

A data collection form was used that details general data of each patient such as age, sex, BMI, comorbidities (Type 2 DM, HTN, Obesity) and complications (Sepsis and Septic Shock) that they presented during hospitalization, as well as ARI. and its corresponding stage.

### STATISTIC ANALYSIS

The multivariate analysis was carried out with the bivariate logistic regression technique and the method = Advance by steps (likelihood ratio) was used to calculate a predictive model of Mortality in Covid-19 patients from exposure to ARI adjusted to variables as SOFA score upon admission to the ICU and serum level of c-reactive protein, obtaining an adequate predictive model (omnibus test: x2: 37.19; p: 0.000), with predictive capacity of 27% (Nagelkerke R squared : 0.272). Likewise, the assumptions of existence of independence between the observations of the dependent and independent variables (Durbin -Watson Test: 1.65) and the non-existence of multicollinearity (Tolerance. 0.92, VIF: 1.08) were validated.

The quantitative variables were analyzed through normal distribution, using the Kolmogorov-Smirnov technique, finding that all these variables have a non-normal distribution (p=0.000), with the exception of age (p0.28) and creatinine at admission, having Keep in mind that non-parametric tests were used to contrast the hypotheses in this study.

### VARIABLES

As the main variable, we used AKI, which is defined as an increase in serum creatinine ≥ 0.3 mg/dl in 48 h or an increase in creatinine ≥ 1.5 times the initial value within 7 days or urinary flow ≤0.5 ml/kg/h. in six hours.^16^

It is classified according to severity into:

- Stage 1: increase in creatinine between 1.5 to 1.9 the initial value.
- Stage 2: increase in creatinine between 2.0 and 2.9 of its initial values.
- Stage 3: increase in creatinine greater than 3 times the initial value.

## RESULTS

### Baseline characteristics of the study populations

200 patients from HAPCII-2 Piura were selected, classifying them into 2 study cohorts, the ARI-exposed cohort was made up of 40 patients and the non-exposed cohort was made up of 160 patients. Regarding the baseline characteristics of the cohorts, they were similar in terms of median BMI (25.3 VS 26.9; P=0.100) and SOFA score upon admission to the Emergency Department (3 vs 5; p=0.690). The cohort exposed to ARI had a higher SOFA score upon admission to the ICU (2 VS 6; P=0.000), a higher serum creatinine level upon admission (0.68% vs 0.84%; p=0.000), a greater range of age (59 vs 64; p=0.029) and lower level of Glomerular filtration rate (107.35 vs 47.2; p=0.000), (Table 01).

**Table 01.** Baseline clinical characteristics of the study cohorts.

| | Cohorts | | U de Mann-Whitney | $p^*$ |
| --- | --- | --- | --- | --- |
|  | No Exposure to Acute Kidney Injury (n:160) | Acute Kidney Injury Exposure (n:40) |  |  |
|  | median | median |  |  |
| Age | 59.00 | 64.00 | 2486.500 | **0.029** |
| Body Mass Index | 25.3500 | 26.9850 | 2662.000 | 0.100 |
| C Reactive Protein | 59.500 | 92.500 | 2564.000 | 0.051 |
| Creatinine on Admission | 0.6850 | 0.8450 | 2037.500 | **0.000** |
| Sofa Score upon Emergency Admission | 3.00 | 5.00 | 3073.50 | 0.690 |
| Sofa score in UCI | 2.00 | 6.00 | 2040.00 | **0.000** |
| Glomerular Filtration | 107.3500 | 47.2000 | 120.000 | **0.000** |

Mann-Whitney test
\*Asymptotic sig. (bilateral)
Grouping variable: Cohorts

Regarding the comorbidities that both cohorts presented at the time of the study. The cohort exposed to ARI presents the same frequency of High Blood Pressure (45% vs 30.6%; p=0.08), Type 2 Diabetes Mellitus (20% vs 26.9; p=0.372), Obesity (22.5% vs 24.5%; p=0.982) than the non-exposed group (Table 02).

**Table 2.**
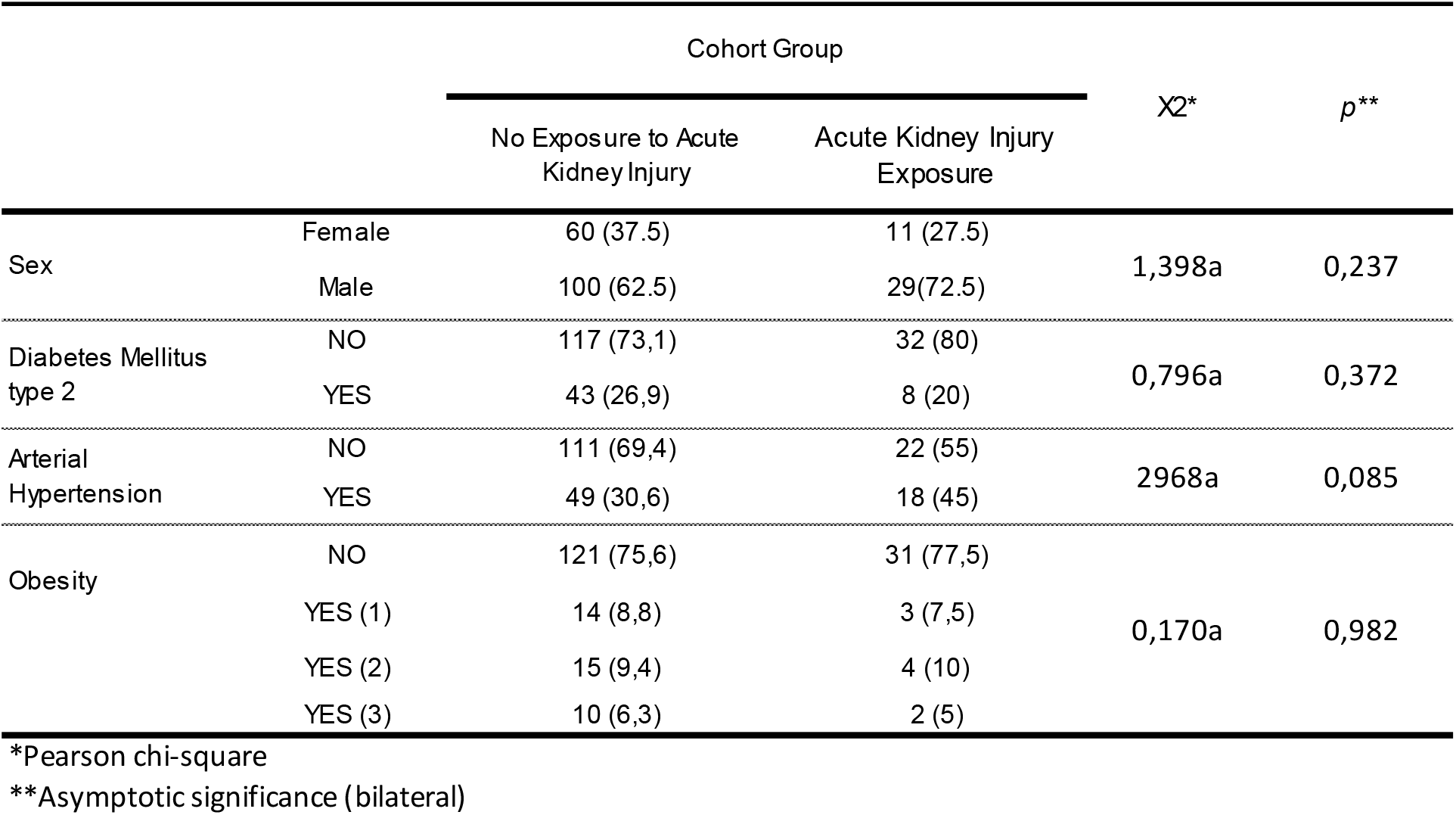
Frequency of metabolic comorbidities in the study cohorts.

|  |  | Cohort Group |  | X<sup>2</sup>* | p<sup>**</sup> |
| --- | --- | --- | --- | --- | --- |
|  |  | No Exposure to Acute Kidney Injury | Acute Kidney Injury Exposure |  |  |
| Sex | Female | 60 (37.5) | 11 (27.5) | 1,398a | 0,237 |
|  | Male | 100 (62.5) | 29(72.5) |  |  |
| Diabetes Mellitus type 2 | NO | 117 (73,1) | 32 (80) | 0,796a | 0,372 |
|  | YES | 43 (26,9) | 8 (20) |  |  |
| Arterial Hypertension | NO | 111 (69,4) | 22 (55) | 2968a | 0,085 |
|  | YES | 49 (30,6) | 18 (45) |  |  |
| Obesity | NO | 121 (75,6) | 31 (77,5) | 0,170a | 0,982 |
|  | YES (1) | 14 (8,8) | 3 (7,5) |  |  |
|  | YES (2) | 15 (9,4) | 4 (10) |  |  |
|  | YES (3) | 10 (6,3) | 2 (5) |  |  |

\*Pearson chi-square
\*\*Asymptotic significance (bilateral)

The frequency of hemodynamic and pulmonary complications was evaluated in the study cohorts during hospitalization, finding that the group exposed to ARI presented a higher frequency of Septic Shock (25% vs 10.6%; p= 0.01). No statistical differences were found between the frequency of hemodynamic complications: Sepsis (100% vs 96.3%), Septic Shock (25% vs 10.6%) and the study cohort (Table 3).

**Table 3.** Frequency of hemodynamic complications during hospitalization in the study cohorts.

|  |  | Cohort Group |  | X<sup>2</sup>* | p<sup>**</sup> |
| --- | --- | --- | --- | --- | --- |
|  |  | No Exposure to Acute Kidney Injury | Acute Kidney Injury Exposure |  |  |
| Sepsis | NO | 6 (3,8) | 0 | 1,546a | 0,214 |
|  | YES | 154 (96,3) | 40 (100) |  |  |
| Septic shock | NO | 143 (89,4) | 30 (75) | 5,663a | 0,017 |
|  | YES | 17 (10,6) | 10 (25) |  |  |

\*Pearson chi-square
\*\*Asymptotic significance (bilateral)

### Mortality, hospital stay and mortality predictors in ARI-exposed cohorts

Mortality was higher in the cohort exposed to ARI (95% vs 71.9%), showing a significant statistical association (p=0.002) and a 5-fold Relative Risk of Death in the patient exposed to ARI compared to the non-COVID 19 patient. exposed to ARI (RR=5.837; CI: 1.46-23.2) (Table 4).

**Table 4.** Relative Risk of Mortality in patients with COVID 19 exposed to ARI during hospitalization.

|  |  | Cohort Group |  | X<sup>2</sup>* | p** | RR | 95% Confidence Interval |  |
| --- | --- | --- | --- | --- | --- | --- | --- | --- |
|  |  | No Exposure to Acute Kidney Injury | Acute Kidney Injury Exposure |  |  |  | Lower | Higher |
| Mortality | Survivors | 45 (28,1) | 2 (5) | 9519a | **0,002** | **5.837** | 1.463 | 23.289 |
|  | Deceased | 115 (71,9) | 38 (95) |  |  |  |  |  |

\*Pearson chi-square
\*\*Asymptotic significance (bilateral)
RR: Relative risk

The cohort of Covid-19 patients exposed to ARI has a longer median hospital stay (17d vs 12.5d), but this dissimilarity in days is not statistically significant (p=0.05). In Graph 01 we can see how the confidence intervals intersect in both groups.

**Graph 01.**
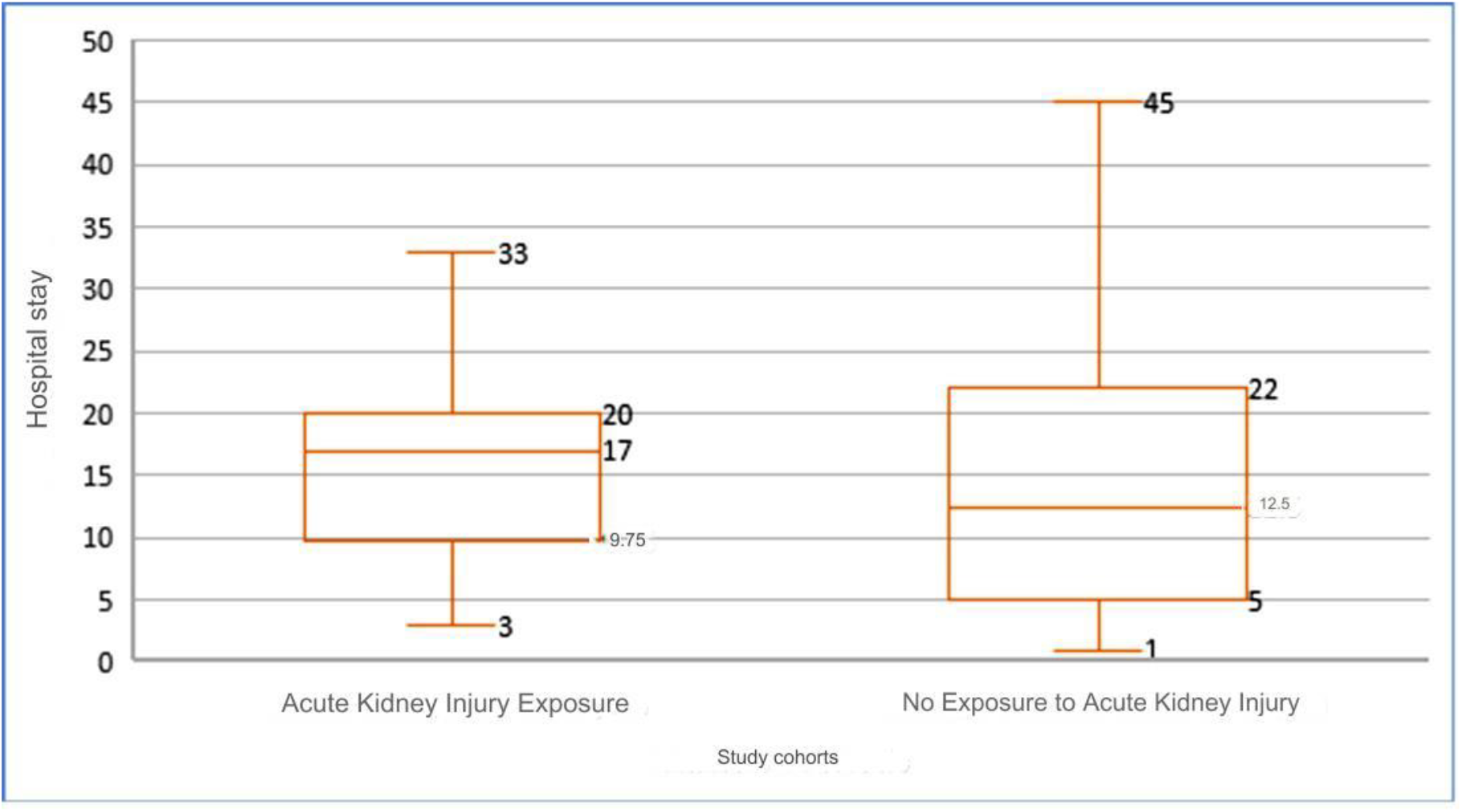
Hospital stay in patients infected with COVID 19 exposed to ARI during hospitalization.

The survivors of 30 days of hospitalization had a SOFA score upon admission to the ICU below 7 points compared to deceased patients who had a SOFA score of up to 14 points. These scores presented a direct correlation with mortality. The mortality prediction was greater than 90% when the SOFA score upon admission to the ICU was greater than 7 points (Graph 02).

**Graph 02.**
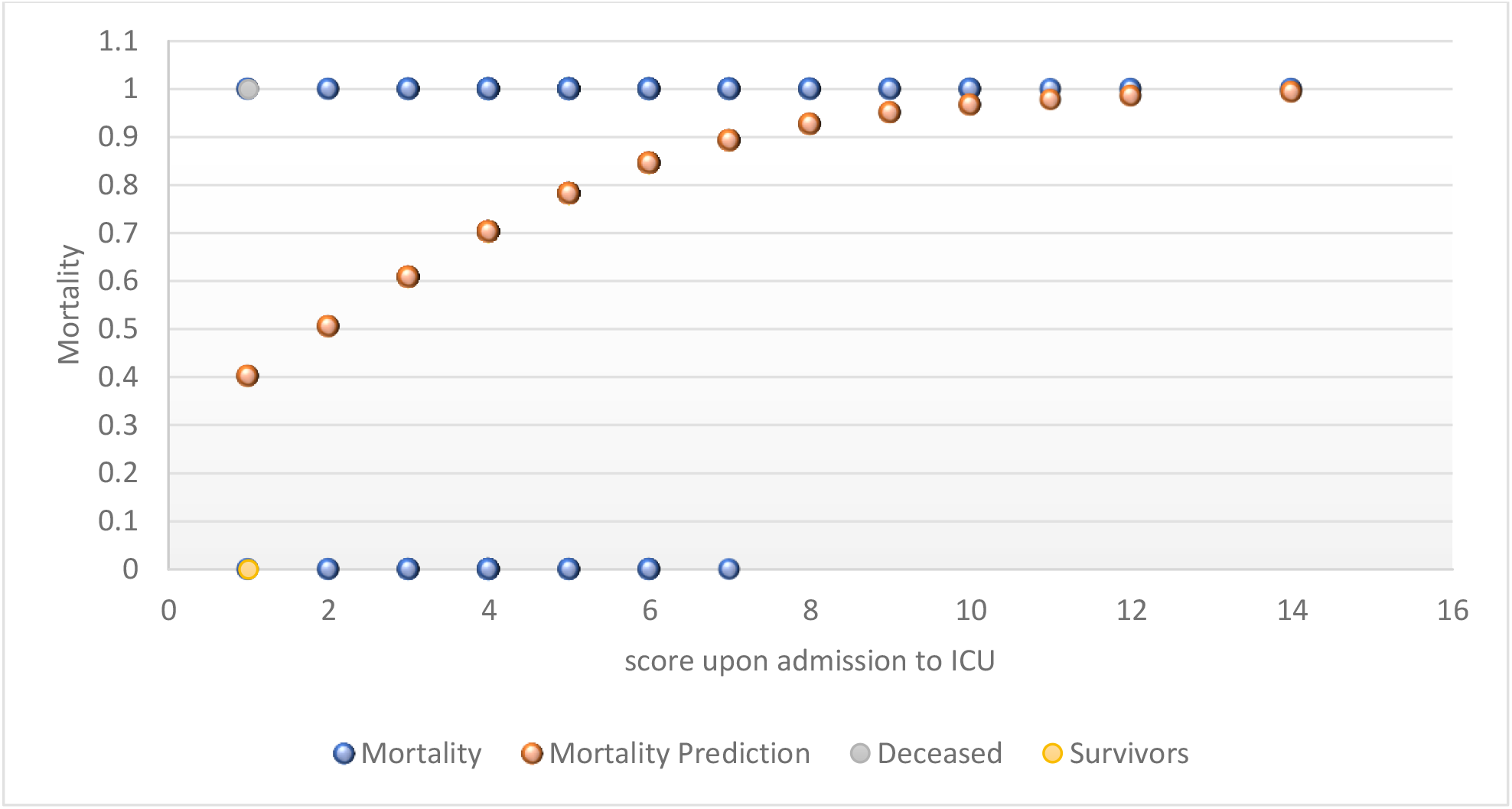
Prediction of mortality in patients diagnosed with COVID 19 based on the SOFA score upon admission to the ICU.

It was shown that: Exposure to ARI during hospitalization increases the risk of death by 5 times compared to COVID patients not exposed to this variable (OR: 5.035; CI: 1.101 – 23.028), when this variable is adjusted to the admission SOFA score to ICU and serum CRP level.

This model also demonstrates that for each point that increases the SOFA score upon admission to the ICU, the risk of mortality increases by 39% (OR: 1.391; CI: 1.103 – 1.754), in addition, each increase in mg/dl of C-Reactive Protein increases the risk of mortality in 2% (OR: 1.021; CI: 1.008 – 1.035), (Table 05).

**Table 05.** Predictive model of mortality in COVID patients from ARI exposure adjusted to variables such as SOFA score upon admission to the ICU and serum level of C-Reactive Protein.

|  | *B* | Sig. | OR | 95% C.I. for EXP(B) |  |
| --- | --- | --- | --- | --- | --- |
|  |  |  |  | Lower | Higher |
| Acute Kidney Injury | 1.616 | 0.037 | 5.035 | 1.101 | 23.028 |
| Sofa score in UCI | 0.330 | 0.005 | 1.391 | 1.103 | 1.754 |
| C Reactive Protein | 0.021 | 0.001 | 1.021 | 1.008 | 1.035 |
| Constant | -1.867 | 0.005 | 0.155 |  |  |

Binary Logistic Regression
Method = Advance by steps (likelihood ratio)

## DISCUSSION

In this research study of 200 patients with severe Covid-19, patients who developed ARI presented an elevated risk of in-hospital mortality compared to the group not exposed to ARI (95% vs 71.9%), thus demonstrating a statistical association significant mortality, with the risk of death being 5 times higher in patients diagnosed with severe Covid-19 exposed to ARI. The results found are similar to those acquired in a study carried out in El Salvador^17^, where it was shown that 52% of patients diagnosed with severe Covid-19 presented ARI compared to 48% who did not develop this complication. In another study carried out by Tarragón et al.^4^ in Spain, a frequency of 22% mortality was found in patients with Covid-19 who developed ARI. Similarly, Li et al.^12^ showed that approximately 44% of patients presented ARF during hospitalization in the ICU, presenting higher in-hospital mortality. Furthermore, in the study by Hamilton et al.^18^, it was found that 20.3% of patients hospitalized for Covid-19 developed ARI during hospitalization, presenting a mortality rate almost 2 times higher compared to patients who did not present ARI (52.4% vs 26.3%). These similar findings between the studies could be due to the renal tropism that the Covid-19 virus presents, described in various mechanisms, one of them being the high affinity of the microorganism for the ACE-2 receptor, which is expressed both at the of lung tissue and in kidney cells, being 100 times greater in the kidneys than in the lung. By increasing the expression of the virus, it blocks ACE-2, causing an increase in the activation of the renin-angiotensin system and the inflammatory response, which helps to exacerbate the disease, and when vascular alterations occur, they trigger hypoxia and hypoperfusion that It generates greater oxidative stress, aggravating lung damage and other organs, causing death.^19^

Another similarity is that the study populations were more men than women, and an association relationship between sex and ARI could develop, however, this has not yet been proven. Furthermore, the patients included in the studies were older, with a median age that varied between 53, 64 and 70 years, in accordance with the median of this study, which was 64 years in patients exposed to ARI. This association could be due because the kidney in elderly patients is more prone to acute stress caused by viral infection, related to the work of T and B cells, and to the overproduction of type 2 cytokines that can cause poor control of viral replication and more proinflammatory responses triggering a poor prognosis.^13^

One of the variables described in this study is the Glomerular Filtration Rate (GFR), which according to Cei et al.^20^ showed a high capacity to detect alterations in renal function in Covid-19 patients and was related to a high incidence of in-hospital mortality when the GFR was lower (<60 ml/min/1.73 m2), similar to what which is shown in our results, where patients who had a low GFR (47.2 ml/min/1.73 m2) presented ARF and subsequently died from it. This could be due to the alteration in the activity of ACE/ACE2 due to the invasion of the virus to the kidney cells, triggering harmful effects at the level of the renal vasculature, deterioration of proximal tubular function and lower glomerular filtration rate as a consequence of hypoperfusion of the kidney. organ.^19^

On the other hand, it was identified that the cohort exposed to ARI had a higher level of serum creatinine at admission compared to the non-exposed cohort (0.68% vs 0.84%), which is similar to the results obtained by Cheng et al^21^, showing that elevated creatinine levels predicted a higher probability of developing ARF during hospitalization. Furthermore, in the study by Gómez-Paz et al.^22^ found that higher creatinine values were related to a higher death rate in Covid-19 patients who developed ARF, which could be due to the loss of renal excretory function, giving way to the storage of nitrogenous products such as urea and creatinine. Furthermore, when there is a decrease in GFR (<50%), creatinine becomes a functional marker, its excretion depending not only on glomerular filtration but also on the renal tubules, reaching 50% as a compensation mechanism by reducing the glomerular filtration rate.^23^

Furthermore, Hirsch et al.^24^ showed that 36.6% of patients who developed ARF showed high rates of associated morbidities such as HBP and Type 2 DM, concluding that these variables could contribute to the development of ARF in patients with Covid-19. In the same way, Oweis et al.^25^, where patients with Covid-19 were elderly and had HTN and Diabetes, concluding that these comorbidities increased the risk of ARF, in opposition to the results of this study, which shows that the frequency of Arterial Hypertension and Diabetes Mellitus were almost the same in the ARI exposed cohort and in the unexposed cohort (45% vs 30.6% and 20% vs 26.9 respectively), which is due to the fact that in our reality, patients have a high incidence of Diabetes Mellitus of 43.8% and a prevalence of arterial hypertension ranging from 20 to 28.7%.^26^

In this research, a multivariate analysis was carried out where it was found that exposure to ARI during hospitalization increases the risk of mortality by 5 times when this variable is adjusted to the SOFA score upon admission to the ICU and the serum CRP level, demonstrating that by Each point that increases the SOFA score upon admission to the ICU increases the risk of mortality by 39% (OR: 1.391; CI: 1.103 – 1.754), in addition, each increase in mg/dl of C-Reactive Protein increases the risk of mortality by 2 % (OR: 1.021; CI: 1.008 – 1.035). These results are compatible with those found in the study by Cui et al.^27^, in which they showed that 18.1% of patients who developed ARF had a higher SOFA score upon admission to the ICU (4.5 ± 2.1) compared to patients who did not develop ARF (2.8 ± 1.4).), triggering a higher mortality rate after adjusting the SOFA Score, which contrasts with the results of this study, where the SOFA score upon admission to the ICU in patients exposed to ARI was 6 points and in the non-exposed cohort it was of 2, showing that the higher the score, the greater the risk of death, reaching 100% with a SOFA score of 14, which demonstrates its high mortality prediction.

Likewise, in the study carried out by De Almeida et al.^28^, it was found that high values of inflammatory markers, including CRP, were associated with the presence of ARF in patients diagnosed with Covid-19, which could be due to the fact that when there is an ongoing viral infection, white blood cells stimulate greater production of C-Reactive Protein through the release of cytokines to counteract the pathogen, making this an associated factor in triggering ARF and thus predicting the severity of the pathology and its possible outcome, as has been identified in others. studies^24^, being similar to the findings of this investigation, where patients who developed ARF had a higher level of C-Reactive Protein compared to the unexposed cohort (92.5 vs 59.5).

This study has limitations such as, being a retrospective observational investigation, it presents inherent biases associated with it, such as the presence of incomplete data, including urinary volume and laboratory tests, especially creatinine tests, or the lack of data in the clinical history, where patients may not have been correctly classified within the exposed cohort, underestimating the incidence of ARI.

On the other hand, the intervening variables could cause confusion bias when determining the death rate of patients, because any of them can contribute to the result, underestimating the frequency of ARI in patients diagnosed with severe Covid-19. Furthermore, since the research work was carried out in a single center, the result of this study could not be determined with certainty.

Despite the limitations, this study presents certain strengths, such as the veracity of the data, for which we have precisely followed the operational definitions stipulated by the KDIGO Guide in the case of ARF, correctly validating the manifestation of ARF in patients. diagnosed with Covid-19. In addition, a multivariate analysis was carried out to estimate the associated comorbidities (HTN, type 2 DM, Obesity), in order to avoid confounding bias, which could alter the results of the present investigation.

It is concluded that the baseline characteristics of the study cohorts showed similarity in terms of BMI, SOFA score upon admission to the emergency room and cardiovascular comorbidities (HTN, type 2 DM, Obesity), they only showed significant differences in the renal studies (glomerular filtration rate, creatinine) and ICU admission SOFA score. The hemodynamic complication that was associated with the cohort exposed to ARI was Septic Shock. The RR of death in patients infected by Covid-19 exposed to ARI is 5 times higher compared to patients who do not manifest this model of kidney failure. Patients with severe Covid-19 exposed to ARI had a longer hospital stay compared to the unexposed cohort. The SOFA score has a direct correlation with mortality and its predictive capacity for mortality is greater than 90% when the SOFA score upon admission to the ICU is greater than 7 points. ARI exposure adjusted to the SOFA score upon admission to the ICU and the serum CRP level during hospitalization increases the risk of death by 5 times in patients exposed to Covid-19 infection.

## Data Availability

All data produced in the present study are available upon reasonable request to the authors.
All data produced in the present work are contained in the manuscript.

## AUTHORS’ CONTRIBUTIONS

ACO designed the study, collected the data, executed the study, and wrote the article. CFA provided significant assistance with the design and development of the study. RSA reviewed the final version of the manuscript and analyzed the data. All authors approved the final version of the manuscript and were accountable for all aspects of the work, including ensuring its accuracy and integrity.

## ACKNOWLEDGMENTS

I thank Dr. Carlos Eduardo Fajardo Arriola and Dr. Raúl Sandoval Ato, for their patience, support and guidance throughout the preparation of this research work.

## Notes

### Competing Interest Statement

The authors have declared no competing interest.

### Funding Statement

This study did not receive any funding

### Author Declarations

The IRB of the Hospital de la Amistad Peru Corea Santa Rosa II gave ethical approval for this work.

